## Supplemental Table 1 for "*HOXA* Amplification Defines a Genetically Distinct Subset of Angiosarcomas"

| Patient characteristics  (36 patient total) | HOXA-amplified (12)  # (%) | Non-HOXA-amp (24)  # (%) | Significance  p-value FE (MV) |
| --- | --- | --- | --- |
| Gender |  |  |  |
| male | 2 (16.7) | 8 (33.3) | ns |
| Female | 10 (83.3) | 16 (66.7) | ns |
| Location |  |  |  |
| HNFS | 2 (16.7) | 8 (33.3) | ns |
| Breast | 8 (66.7) | 11 (45.8) | ns |
| intrathoracic/abd | 2 (16.7) | 4 (16.7) | ns |
| limb | 0 (0) | 1 (4.2) | ns |
| Prior Radiation |  |  |  |
| yes | 2 (16.7) | 5 (20.8) | ns |
| Age |  |  |  |
| ≤30 years | 1 (8.3) | 4 (16.7) | ns |
| 31-40 years | 4 (33.3) | 8 (33.3) | ns |
| 41-50 years | 0 | 2 (8.3) | ns |
| 51-60 years | 2 (16.7) | 4 (16.7) | ns |
| 61-70 years | 4 (33.3) | 4 (16.7) | ns |
| >70 years | 1 (8.3) | 2 (8.3) | ns |
| Pathology |  |  |  |
| vasoformative | 10 (83.3) | 24 (100) | ns |
| epitheloid | 7 (58.3) | 8 (33.3) | ns |
| Spindel cell | 6 (50) | 17 (70.8) | ns |
| Mutation buRDEN |  |  |  |
| >200 mutations | 1 (8.3) | 7 (29.2) | ns |
| Amplifications |  |  |  |
| CD36 | **8 (66.7)** | **2 (8.3)** | **5.59E-4 (1.62E-2)** |
| KDR | **9 (75)** | **4 (16.7)** | **1.07E-3 (3.11E-2)** |
| PHF1 | 9(75) | 10 (41.7) | ns |
| Mutations |  |  |  |
| p53 | 4 (33.3) | 7 (29.2) | ns |
| KDR | 3 (25) | 5 (20.8) | ns |
| PIK3CA | 2 (16.7) | 4 (16.7) | ns |
| NRAS | 0 | 3 (12.5) | ns |
| HRAS | 1 (8.3) | 1 (4.1) | ns |
| BRAF | 0 | 2 (8.3) | ns |
| RAF1 | 0 | 1 (4.1) | ns |
| NF1 | 2 (16.7) | 2 (8.3) | ns |
| MAP3K13 | 0 | 1 (4.1) | ns |

**Supplemental Table 1:** Patient clinical characteristics and recurrent amplifications/mutation of *HOXA*-amplified and non-*HOXA* amplified angiosarcomas. *HOXA*-amplifications were significantly associated with amplifications of *KDR* and *CD36*.
